# Disease-specific ACMG/AMP guidelines improve sequence variant interpretation for hearing loss

**DOI:** 10.1101/2021.01.27.21250652

**Authors:** Mayher J Patel, Marina T DiStefano, Andrea M Oza, Madeline Y Hughes, Emma H Wilcox, Sarah E Hemphill, Brandon J Cushman, Andrew R Grant, Rebecca K Siegert, Jun Shen, Alex Chapin, Nicole J Boczek, Lisa A Schimmenti, Kiyomitsu Nara, Margaret Kenna, Hela Azaiez, Kevin T Booth, Andrew Griffith, Karen B Avraham, Hannie Kremer, Heidi L Rehm, Sami S Amr, Ahmad N Abou Tayoun, ClinGen Hearing Loss Clinical Domain Working Group

## Abstract

**Purpose:** The ClinGen Variant Curation Expert Panels (VCEPS) provide disease-specific rules for accurate variant interpretation. Using hearing loss-specific American College of Medical Genetics/Association for Molecular Pathology (HL-specific ACMG/AMP) guidelines, the ClinGen Hearing Loss VCEP (HL VCEP) illustrates the utility of expert specifications in resolving conflicting variant interpretations.

**Methods:** A total of 157 variants across nine hearing loss genes were curated and submitted to ClinVar by the HL VCEP. The curation process consisted of collecting published and unpublished data for each variant by biocurators, followed by bi-monthly meetings of an expert curation subgroup that reviewed all evidence and applied the HL-specific ACMG/AMP guidelines to reach a final classification.

**Results:** Before expert curation, 75% (117/157) of variants had single or multiple VUS submissions (17/157) or had conflicting interpretations in ClinVar (100/157). After applying the HL-specific ACMG/AMP guidelines, 24% (4/17) of VUS variants and 69% (69/100) of discordant variants were resolved into Benign (B), Likely Benign (LB), Likely Pathogenic (LP), or Pathogenic (P). Overall, 70% (109/157) variants had unambiguous classifications (B, LB, LP, P). We quantify the contribution of the HL-specified ACMG/AMP codes to variant interpretation.

**Conclusion:** Expert specification and application of the HL-specific ACMG/AMP guidelines effectively resolved discordant interpretations in ClinVar. This study supports the utility of ClinGen VCEPs in helping the community move towards more consistent variant interpretations, which will improve the care of patients with genetic disorders.

## Introduction

Hearing loss (HL) is the most common congenital sensory condition, with approximately 50% of affected individuals having an identifiable genetic etiology^1^. Genetic HL is a heterogeneous condition with more than 100 genes underlying nonsyndromic HL and over 400 genes implicated in syndromic forms of deafness^2,3^. A clinical genetics evaluation is recommended as part of a standard of care diagnostic work-up, and results from genetic testing can often inform clinical management, especially if a genetic syndrome is identified before the onset of additional clinical manifestations^1,4^. As an expert panel of the Clinical Genome Resource (ClinGen), a National Institute of Health funded resource focused on defining the clinical validity of gene and variant contributions to disease, the Hearing Loss Clinical Domain Working Group (HL-CDWG) (https://www.clinicalgenome.org/working-groups/clinical-domain/hearing-loss/) has worked to evaluate gene-disease relationships and standardize variant interpretation in hereditary HL^5,6,7^.

Interpretation of the clinical significance of variants is a rigorous process that involves collating and analyzing available literature and evidence, followed by a formal classification based on this evidence. The ClinVar database (https://www.ncbi.nlm.nih.gov/clinvar/) maintained by the National Center for Biotechnology Information, archives and aggregates variant interpretations from various submitters and indicates whether the submitted interpretations are concordant or discordant. Data sharing through ClinVar provides an invaluable opportunity to identify classification differences and to collaborate with submitters to resolve these discrepancies. A pilot study conducted by four clinical laboratories showed that 87.2% (211/242) of discordant variants were resolved when the variant was reassessed with current criteria and/or through internal data sharing^8^. The resolution of conflicting interpretations helps provide crucial diagnostic information to clinicians and patients.

Here we document the curation efforts of the ClinGen Hearing Loss Variant Curation Expert Panel (HL VCEP) and highlight the successful effort to resolve discrepancies in variant interpretations. The HL VCEP applied the hearing loss-specific (HL-specific) ACMG/AMP guidelines^5^ to interpret the clinical significance of variants in nine genes associated with nonsyndromic or syndromic HL. Our data support the role of ClinGen Variant Curation Expert Panels and the application of disease-specific American College of Medical Genetics/Association for Molecular Pathology (ACMG/AMP) guidelines in resolving discordant variant interpretations present in ClinVar.

## Methods

### HL-CDWG Organizational Structure

The HL-CDWG has worked to create a standardized, thorough set of genes and variants that are associated with syndromic and nonsyndromic HL^5,9^. The members include otolaryngologists, clinical geneticists, molecular geneticists, ClinGen biocurators, clinical researchers, and genetic counselors from 11 different countries including China, France, Israel, Japan, Netherlands, Singapore, South Korea, Spain, Tunisia, the United Arab Emirates, and the United States, representing 26 institutions. In order to accomplish this goal, the HL-CDWG formed two other groups defined by ClinGen as a Gene Curation Expert Panel (GCEP) and a Variant Curation Expert Panel (VCEP).

### HL VCEP Rule Specification and Curation Process

A smaller task team of the HL VCEP worked to provide expert guidance for standardized variant interpretation and adapted the ACMG/AMP guidelines for the interpretation of sequence variants in nine HL genes, specifically *USH2A, SLC26A4, GJB2, MYO7A, CDH23, TECTA, COCH, KCNQ4*, and *MYO6*^5^.

The HL VCEP group then continued to curate variants in these nine genes according to the new HL specific standards with an emphasis on those that have conflicting interpretations in ClinVar. The HL VCEP utilizes monthly meetings in addition to email correspondence to present, review, and reach consensus on the classification of these variants. Each variant was assigned to a single trained ClinGen biocurator who utilized various literature search engines and databases including PubMed, the Deafness Variation Database (DVD)^10^, Human Gene Mutation Database (HGMD), Google Scholar, Leiden Open Variation Database (LOVD), and LitVar to find publications associated with these variants. HL VCEP members also contributed case observations and phenotypic information from their laboratories or clinics. Additionally, ClinVar submitters were contacted via email to provide internal genotype and phenotype data for their submissions so that biocurators could aggregate all the relevant data. Curators utilized ClinGen’s Variant Curation Interface (https://curation.clinicalgenome.org/) to assess and document the applicable rules for each variant. Provisional classifications made by the curators were comprehensively presented to the chairs and members of the HL VCEP during the monthly meetings. Members provided verbal feedback during these calls to help modify and approve the various ACMG/AMP codes as necessary. Monthly calls between the biocurators and the chairs of the group were utilized to finalize the classification of variants and to discuss the language of the classification summary text. The average curation time for an individual variant was 40 minutes. The expert classifications of these variants were submitted to ClinVar on a quarterly basis (**Figure 1**).

**Figure 1.**
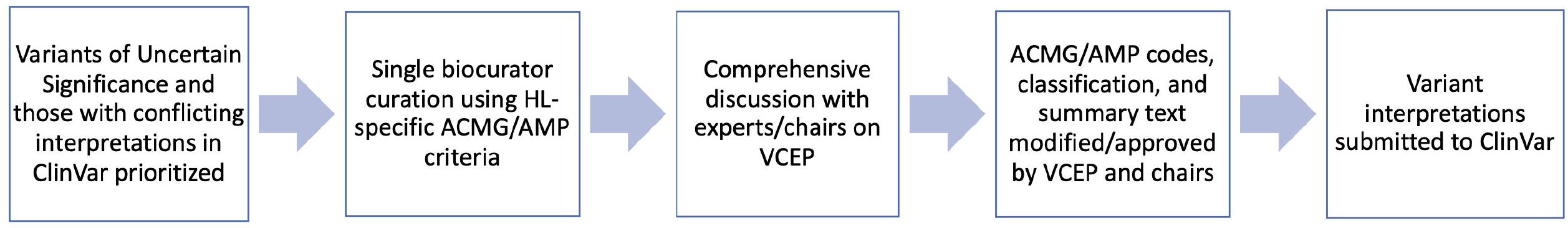
Workflow highlighting the steps in the Expert curation process.\

## Results

### Curated Variants

Since the specification and promulgation of the ACMG/AMP variant rules by the HL VCEP (Oza 2018; Abou Tayoun 2018), 157 variants across nine HL genes (**Supplementary Figure 1, Supplementary Table 1**) underwent an expert review and classification process as shown in **Figure 1**. The majority of variants (79%) were missense (n=124), while the remaining (21%) were in essential (+/−1,2) splice sites (n=4), frameshift (n=7), synonymous (n=5), non-essential splice regions (n=10), nonsense (n=3), inframe deletion (n=1), exon deletion (n=1), start loss (n=1), or UTR (n=1) variants (**Supplementary Table 1**). As of January 8, 2021, 937 classifications have been submitted from 70 submitters for these 157 variants.

Prior to expert curation, 100 variants had conflicting interpretations, 17 variants had VUS classifications, and 40 variants had no conflicts in ClinVar. Of the 100 variants with conflicting interpretations, 76 variants had medically significant conflicting classifications (P/LP vs. VUS/LB/B as defined by Harrison et al 2017^8^) and 24 variants had VUS vs LB/B conflicts. Only three variants had conflicting classifications of P/LP vs LB/B. For the variants with no conflicting classifications, 33 variants were supported by multiple submissions, five variants had only one submission, and two variants had no previous submissions. Of these 40 variants, some were chosen to pilot the initial specifications of the rules while others were considered clinically significant variants or common pathogenic variants in a specific subpopulation^5^ (**Supplementary Figure 1**).

### Expert Curation Outcomes

After applying the HL-specific ACMG/AMP variant rules, 73 out of the 117 variants (63%) with either conflicting or VUS classifications, were classified as B, LB, LP, or P, while 44 variants (37%) were classified as VUS (**Figure 2**). The majority of the resolved classifications belonged to the VUS vs LB/B conflicting category where 20 out of those 24 variants (83%) were classified as B or LB and the remaining as VUS. On the other hand, 38 out of the 63 variants (60.3%) with VUS vs LP/P conflicting interpretations were classified as P or LP, one was classified as LB, and the remaining to VUS. Four out of 17 variants (24%) with single or multiple VUS submissions were classified as B (n=1) or LB (n=2) or LP (n=1). Of the 40 non-VUS variants without conflicts, 13 (32.5%) were classified as B or LB, while 23 (57.5%) were classified as P or LP. Overall, 109 out of the 157 variants (69%) were assigned non-VUS classifications (B, LB, LP, P) after applying the HL-specific ACMG/AMP modified variant interpretation guidelines by the expert group (**Figure 2**).

**Figure 2.**
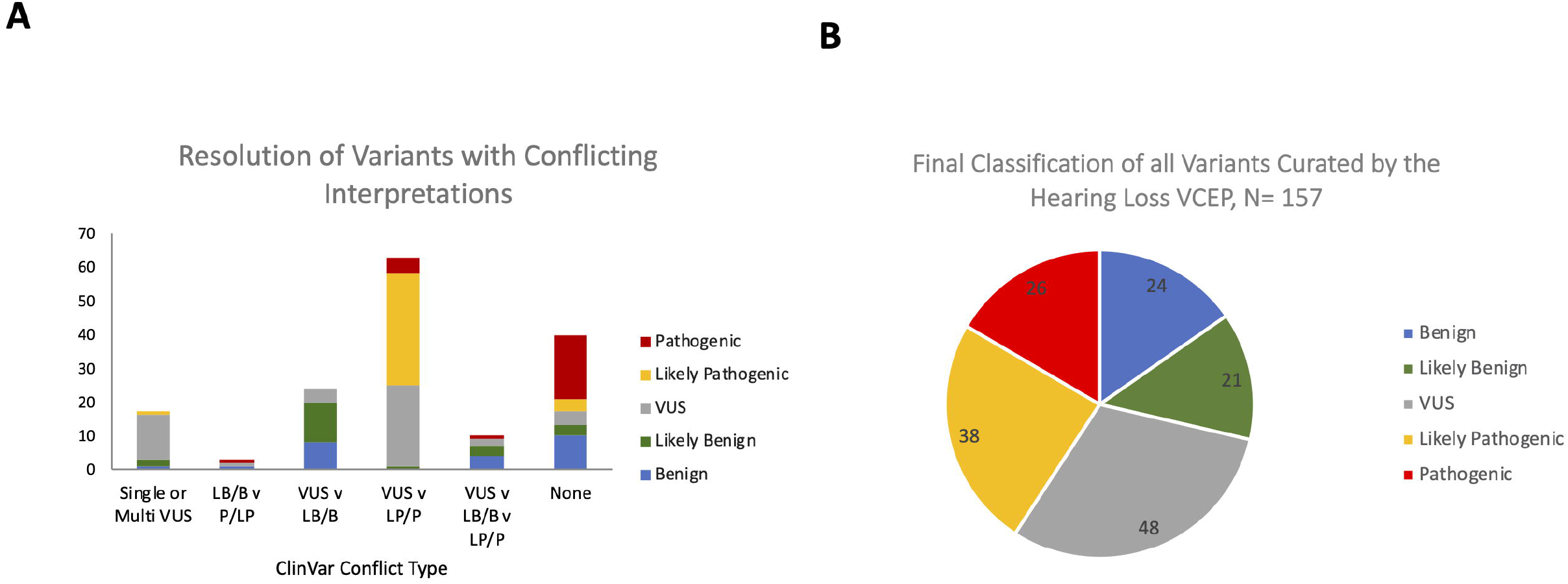
Outcomes of expert variant interpretation. *A*, Resolution of variants with different conflict types into Pathogenic, Likely Pathogenic, VUS, Likely Benign, or Benign by the ClinGen HL EP. Y-axis represents the number of variants; X-axis represents the conflict type in ClinVar. *B*, Final classifications of all variants curated by the Hearing Loss VCEP (N=157).

### Criteria contribution to the final classification

In classifying the 157 variants, the most commonly applied criteria codes were PM2 (absence or rare occurrence in population databases) at two different strength levels (PM2_Supporting and PM2) and PM3 (allelic observations in recessive cases) at four different strength levels (PM3_Supporting, PM3, PM3_Strong, PM3_Very Strong), as specified by the HL VCEP (Oza et al., 2018). Both the PM2 and PM3 codes were used 80 times and were applied to 77% (49/64) and 88% (56/64) of the variants classified as P/LP, respectively (**Supplementary Figure 2**). The in silico prediction code PP3, specified using REVEL^5,11^, was applied 64 times as the second most commonly used code, and was applied to 59% (38/64) of P/LP variants. The BA1, BS1, and BS1_Supporting allele frequency codes, as specified by the expert panel, were applied 59 times and to 91% (41/45) of the B/LB classifications. The phenotype code PP4 was used 47 times and was applied to 58% (37/64) of P/LP variants. The specified segregation code PP1 at three different strength levels (PP1, PP1_Moderate, PP1_Strong) was used 31 times and was applied to 41% (26/64) of P/LP variants. Several other codes were utilized as specified by the HL VCEP and as shown in **Supplementary Figure 2**.

## Discussion

Our study highlights the immense value of disease-specific ACMG/AMP guidelines for accurate variant interpretation. For genetic HL, 25 out of the 28 ACMG/AMP criteria were specifically tailored to hereditary HL^5,9^. The HL VCEP included an international group of scientists, clinicians, geneticists, genetic counselors, and laboratory directors working in the auditory field. This panel leveraged the collective members’ expertise and knowledge, along with internal unpublished datasets accessible to the group, to characterize several HL attributes such as prevalence, penetrance, genetic and allelic heterogeneity, and phenotypic presentations (onset, severity, frequencies affected, symmetry and audiometric profile) specific to several HL-associated genes. This information was then used to define the criteria for variant classification and to resolve conflicting classifications.

Our panel implemented these HL-specific guidelines to classify 157 variants. One hundred and nine variants (69%) were classified into non-VUS categories. Interestingly, the commonly used classification criteria codes were either specified for HL (e.g. PM2, BA1, BS1, BS1_Supporting, PP3, PP4), or, in collaboration with the ClinGen Sequence Variant Interpretation (SVI) workgroup, specified more broadly for recessive disorders (e.g. PM3) or any disorder (e.g. PP1). Application of criteria without disease-specific guidance is a large contributor of conflicting classifications, and emphasizes the importance of expert guidance and specification in usage of the ACMG/AMP rules. Several variants classified as P and LP also had BA1 and BS1 codes that were applied to them. Given that many of these variants were founder variants in specific subpopulations or had other evidence such as case-control data and case counts, the group decided to exclude the conflicting benign population frequency codes from impacting the overall clinical interpretation of the variant.

Sharing of internal clinical laboratory case data among members of the HL VCEP has also served as an invaluable method for resolving variants with conflicting interpretations. For example, the c.1708G>A (p.Val570Ile) variant in *SLC26A4* previously had a conflicting classification and a medically significant difference of VUS vs LP. The HL VCEP resolved this conflict by counting 2 compound heterozygous internal cases from the Laboratory of Molecular Medicine at Mass General Brigham Personalized Medicine, which led to the application of PM3_Strong, PP4, and a LP classification. Without these internal cases, the variant would have been classified as VUS due to a lack of published case-level evidence and a highly-specific phenotype.

Our curation process included bimonthly calls where biocurators first present all evidence used for each variant classification to the experts. Based on the feedback, biocurators often reached out to ClinVar submitters, authors of certain publications, and/or any of the experts to gather additional unpublished data required for final variant interpretation. The final evidence is then discussed on intervening monthly calls with the HL VCEP chairs, and if needed on subsequent calls with the larger group, to reach a final classification, including a text-based variant evidence summary documenting all applied evidence to be submitted to ClinVar and publically accessible through ClinGen’s Evidence Repository (https://erepo.clinicalgenome.org/evrepo/). This process has been, and continues to be, optimized for more efficient expert interpretations while still leveraging the diverse expertise within the group. It is important to note that this initial effort was mostly focused on variants with conflicting or uncertain significance to highlight the utility of the specified rules. In this process, a significant effort was dedicated to contacting ClinVar submitters and obtaining additional unpublished evidence to resolve interpretation discrepancies.

We have focused this pilot project on variants in nine HL-associated genes, *USH2A, SLC26A4, GJB2, MYO7A, CDH23, TECTA, COCH, KCNQ4*, and *MYO6*, for which ACMG/AMP specifications were made. However, most of those specified rules are equally applicable to other HL-associated genes. As such, the population frequency (BA1, BS1, PM2), allelic (PM3, BP2), *de novo* (PS2, PM6), segregation (PP1), predicted effects (PVS1, PM4, PP3, BP3, BP4, BP7), case-control (PS4), and their varied strength levels as specified by the expert group, could be extended to other genes. The HL VCEP is in the process of expanding the genes for which the current modified guidelines could be used. The group will also continue to prioritize curation of variants in HL-genes with medically significant differences and those of uncertain significance.

In summary, our study shows that expert variant curation using disease-specific modifications of ACMG/AMP guidelines resolves discrepancies in variant classification, leading to more consistent results for patients in need of accurate diagnoses for treatment and management decisions.

## Data Availability

All Data has been submitted to ClinVar. Please refer to Supplementary table 1 for a list of variants and their ClinVar IDs

## Data Availability

All variants were submitted to ClinVar by the HL-EP. Refer to Supplementary Table 1 for a list of all variants and their ClinVar IDs.

## Acknowledgements

The authors would like to acknowledge Larry Babb for his help in compiling data from ClinVar to conduct some of these analyses and additional members of the Hearing Loss Clinical Domain Working Group including Sonia Abdelhak, John Alexander, Zippora Brownstein, Rachel Burt, Byung Yoon Choi, Lilian Downie,Thomas Friedman, Anne Giersch, John Greinwald, Jeffrey Holt, Makoto Hosoya, Un-Kyung Kim, Ian Krantz, Suzanne Leal, Saber Masmoudi, Tatsuo Matsunaga, Matías Morín, Cynthia Morton, Hideki Mutai, Arti Pandya, Richard Smith, Mustafa Tekin, Shin-Ichi Usami, Guy Van Camp, Kazuki Yamazawa, Hui-Jun Yuan, Elizabeth Black-Zeigelbein, and Keijan Zhang.

## Author Information

Conceptualization: AAT, SA, HLR; Data curation: MJP, MTD, AMO, MYH, EHW, SHE, BJC, RG, RKS, AC; Formal Analysis: AAT, MJP; Funding acquisition: HLR; Writing – original draft: AAT, MJP; Writing – review & editing: MTD, AMO, MYH, EHW, SHE, BJC, RG, RKS, AC, JS, NJB, LAS, KN, MK, HA, KTB, AG, KBA, HK, HLR, SA.

## Ethics Declaration

Not applicable

## Figure Legends

**Supplementary Figure 1**. *A*, Distribution of all 157 variants by conflict type. *B*, Distribution of variant by gene.

**Supplementary Figure 2**. Usage (A) and contribution (B) of the different ACMG/AMP codes, specified by the HL EP, to the final variant classification.

