## Supplementary figures and images for "Disease-specific ACMG/AMP guidelines improve sequence variant interpretation for hearing loss"

### Supplementary Figure 1

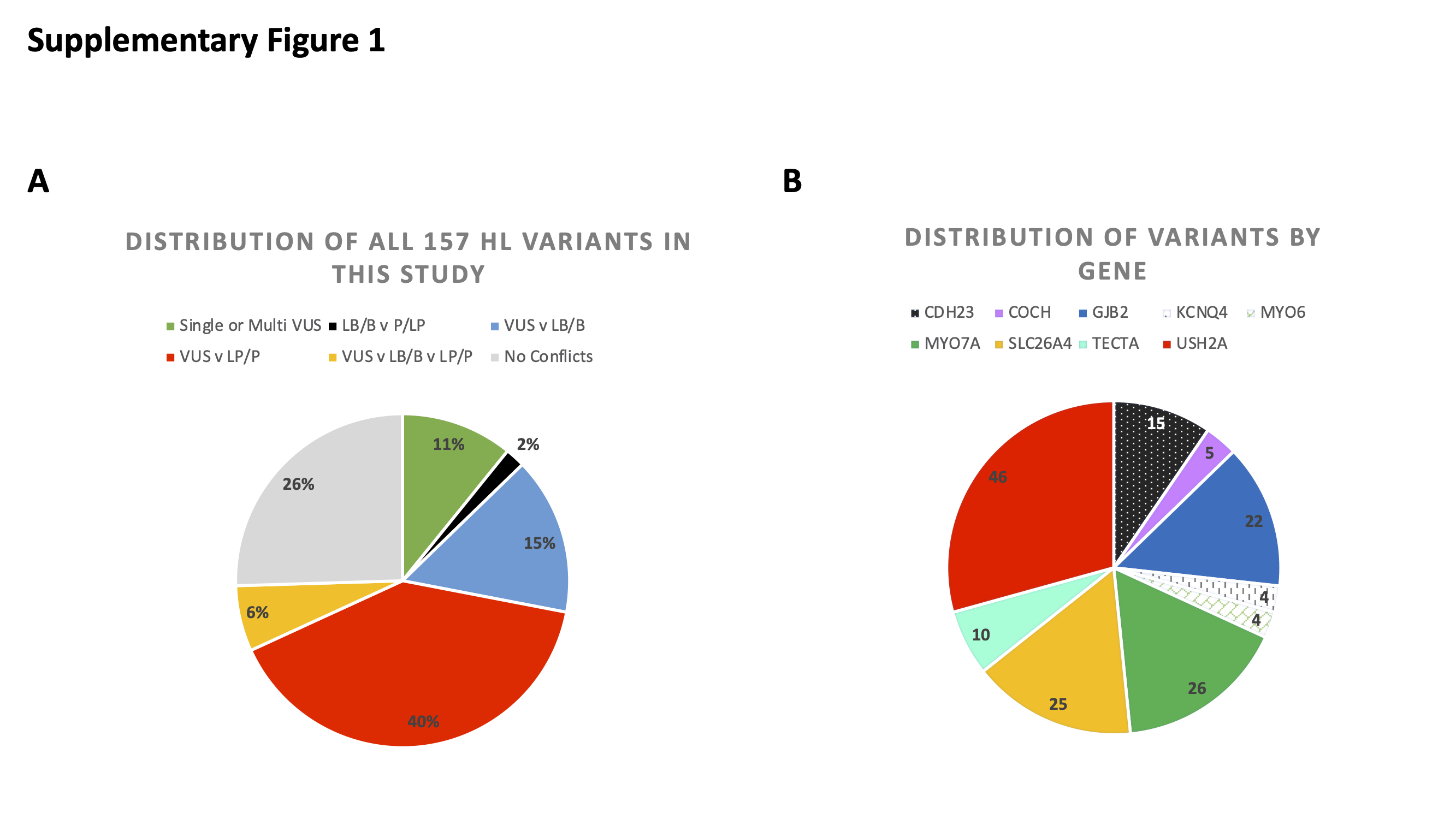

### Supplementary Figure 2

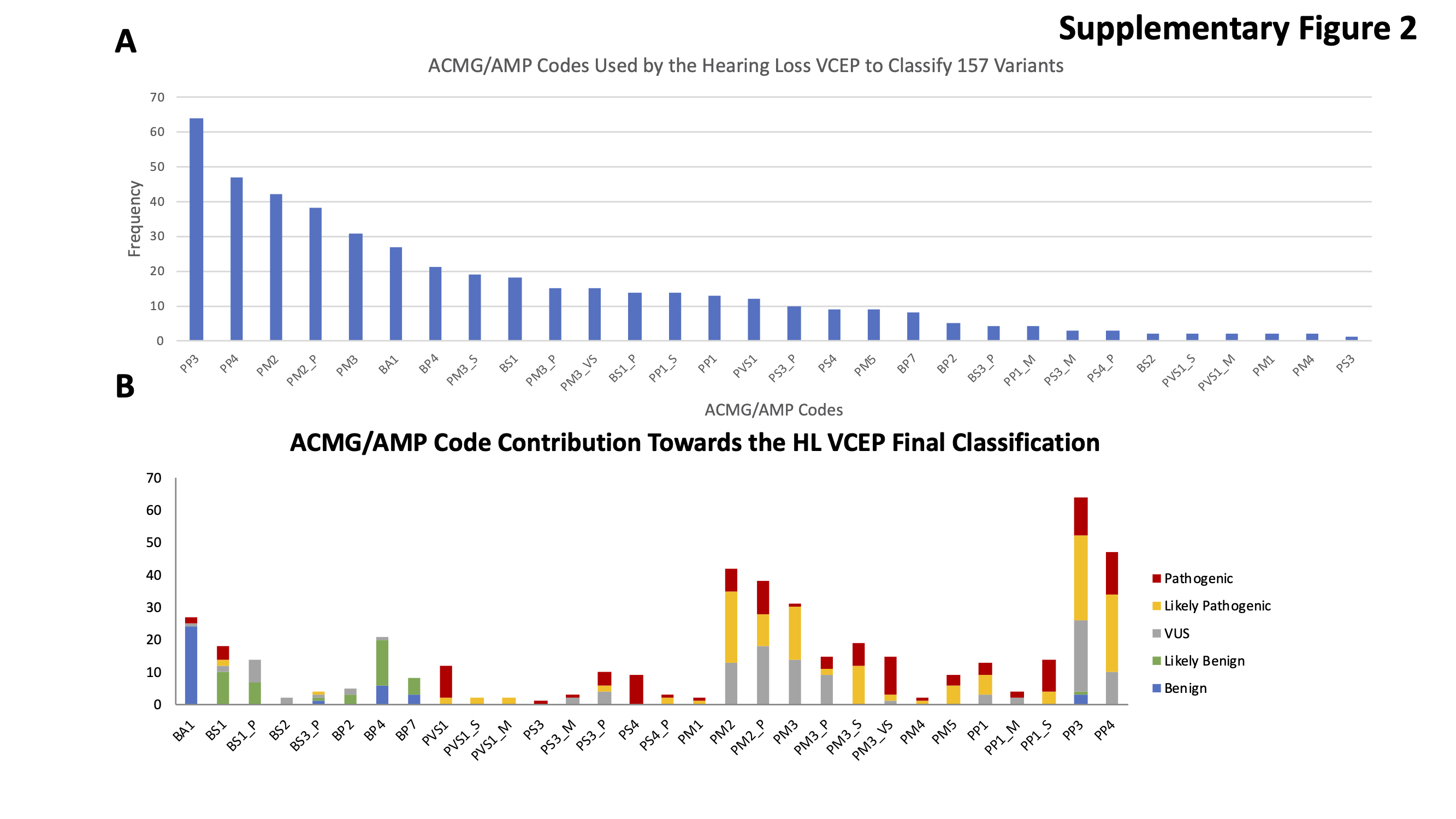
